# Unified real-time environmental-epidemiological data for multiscale modeling of the COVID-19 pandemic

**DOI:** 10.1101/2021.05.05.21256712

**Authors:** Hamada S. Badr, Benjamin F. Zaitchik, Gaige H. Kerr, Nhat-Lan H. Nguyen, Yen-Ting Chen, Patrick Hinson, Josh M. Colston, Margaret N. Kosek, Ensheng Dong, Hongru Du, Maximilian Marshall, Kristen Nixon, Arash Mohegh, Daniel L. Goldberg, Susan C. Anenberg, Lauren M. Gardner

## Abstract

An impressive number of COVID-19 data catalogs exist. None, however, are optimized for data science applications, *e.g*., inconsistent naming and data conventions, uneven quality control, and lack of alignment between disease data and potential predictors pose barriers to robust modeling and analysis. To address this gap, we generated a unified dataset that integrates and implements quality checks of the data from numerous leading sources of COVID-19 epidemiological and environmental data. We use a globally consistent hierarchy of administrative units to facilitate analysis within and across countries. The dataset applies this unified hierarchy to align COVID-19 case data with a number of other data types relevant to understanding and predicting COVID-19 risk, including hydrometeorological data, air quality, information on COVID-19 control policies, and key demographic characteristics.

## Background & Summary

The ongoing COVID-19 pandemic has caused widespread illness, loss of life, and societal upheaval across the globe. As the public health crisis continues, there is both an urgent need and a unique opportunity to track and characterize the spread of the virus and sensitivity of disease transmission to demographic, geographic, socio-political, seasonal and environmental factors, including influence of climate and air quality conditions. The global research and data science communities have responded to this challenge with a wide array of efforts to collect, catalog, and disseminate data on COVID case numbers, hospitalizations, mortality, and other indicators of COVID incidence and burden.^1-12^ Some of these efforts have attempted to integrate data at regional to global scale, including inventories at the finest geographic scale available. While these databases have supported a tremendous volume of research, risk monitoring, and public discussion, they have limitations that may have slowed research progress and, at times, allowed for the production of spurious results. Even the best inventories suffer from the challenge of managing and sharing large volumes of data of inconsistent frequency, resolution, and quality, and most public facing databases do not include data consistency checks that can be critical for data science applications. A unified dataset will help in accelerating multiscale spatiotemporal modeling by eliminating the extra time-consuming steps needed to clean, standardize, and merge the different data sources.

Recognizing the need to: (1) harmonize naming and coding conventions, (2) implement quality control for COVID-19 case counts of different types, and (3) systematically align potential predictors with COVID-19 data, our *Unified COVID-19 Dataset* harmonizes COVID-19 data from credible data sources at multiple administrative levels. The dataset maps all geospatial units globally into a unique identifier, standardizing administrative names, codes, dates, data types, and formats and unifying variable names, types, and categories as well as curating the data and fixing confusing entries that arise from the conflicting names of the same geographic units, different reporting strategies and schedules, and accumulation of case counts. The dataset is optimized for machine learning applications and includes key components for epidemiology, including demography, hydrometeorology, air quality, policy, and healthcare accessibility. Most components are updated on a daily basis while time-consuming data extraction for hydrometeorological variables, with and without population weighting, are updated weekly. The dataset is disseminated through the Center for Systems Science and Engineering (CSSE) at Johns Hopkins University (JHU), the source of the widely accessed JHU Coronavirus Dashboard.^1^

## Methods

We collect epidemiological data from different sources, translate the data records, and check the available case types. Then, the variable and unit names are standardized and geo-coded using a unified geospatial identifier (ID) to support aggregation at different administrative levels and consistent merging into a single time-varying epidemiological dataset file. The case types that are not included in the raw data are derived from the existing case types whenever possible (e.g., deriving active cases from confirmed cases, recoveries, and deaths). A lookup table provides key geographic names and codes while the static data fields, including air quality estimates, are combined in a separate dataset file. Time-varying hydrometeorological and policy data are processed to extract the variables and indices for each geospatial ID at a daily resolution.

### Geospatial ID

The spatial coverage of the dataset is shown in the world map in **Figure 1** and the geospatial ID system is shown in **Figure 2**. The national-level IDs are based on ISO 3166-1 alpha-2 codes. The subnational administrative levels for the United States (at the state and county levels) are based on the Federal Information Processing Standard (FIPS) codes. For Europe, all administrative levels use the Nomenclature of Territorial Units for Statistics (NUTS) codes. Globally, the principal subdivisions (e.g., provinces or states) use ISO 3166-2 code while higher resolution units are based on local identifiers (e.g., for Brazil, municipalities use IBGE codes from the Brazilian Institute of Geography and Statistics).

**Figure 1:**
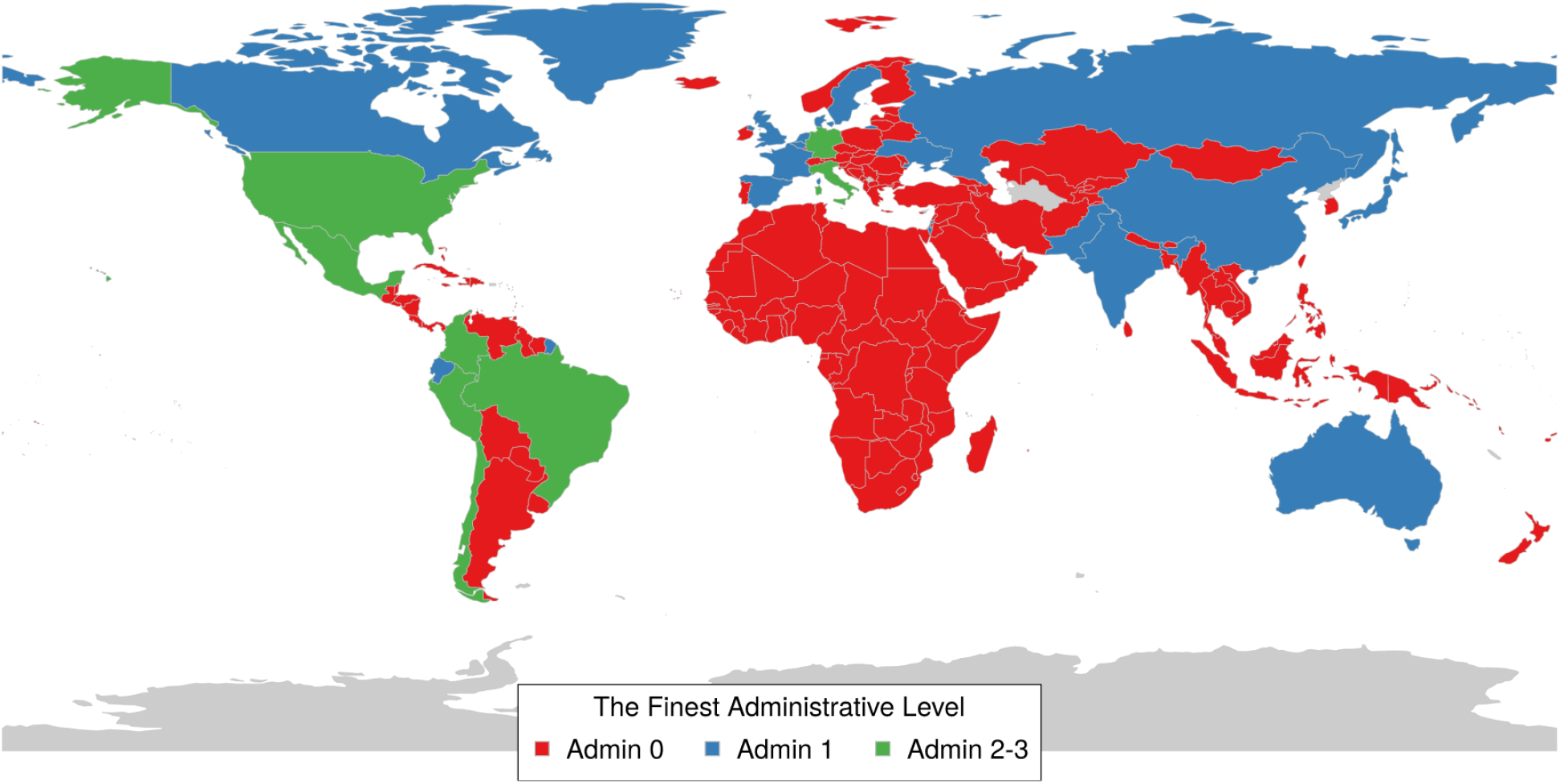
Spatial coverage map for the unified COVID-19 dataset.

**Figure 2:**
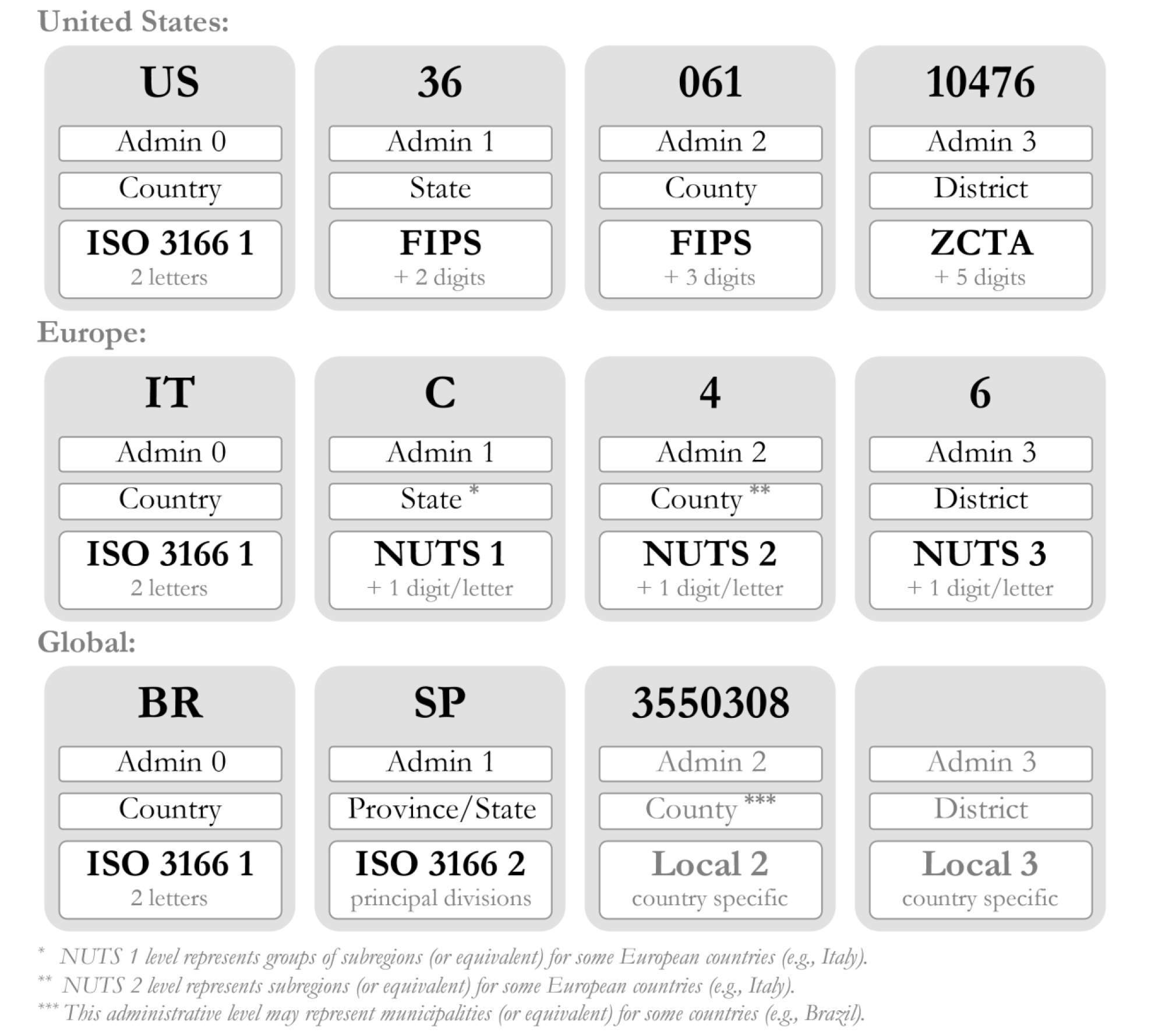
Geospatial ID used for the unified COVID-19 dataset.

### Population Weighting

Population weighting is applied to gridded environmental data (hydrometeorology and air quality) to account for variation in the spatial distribution of the exposed human population within each unit. Gridded Population of the World v4 (GPWv4) population count data with adjustment to match United Nations estimates are obtained from the Center for International Earth Science Information Network (CIESIN) Socioeconomic Data and Applications Center SEDAC.^13^ These counts are then applied as weights by calculating the fraction of the population within each unit at each level of the administrative hierarchy contained in each grid cell, multiplying gridded environmental variables by this fraction, and summing for the administrative unit.

## Data Records

**Table 1** summarizes the lookup table keys with the different unit IDs, names, codes, centroid coordinates, and population.

**Table 1:**
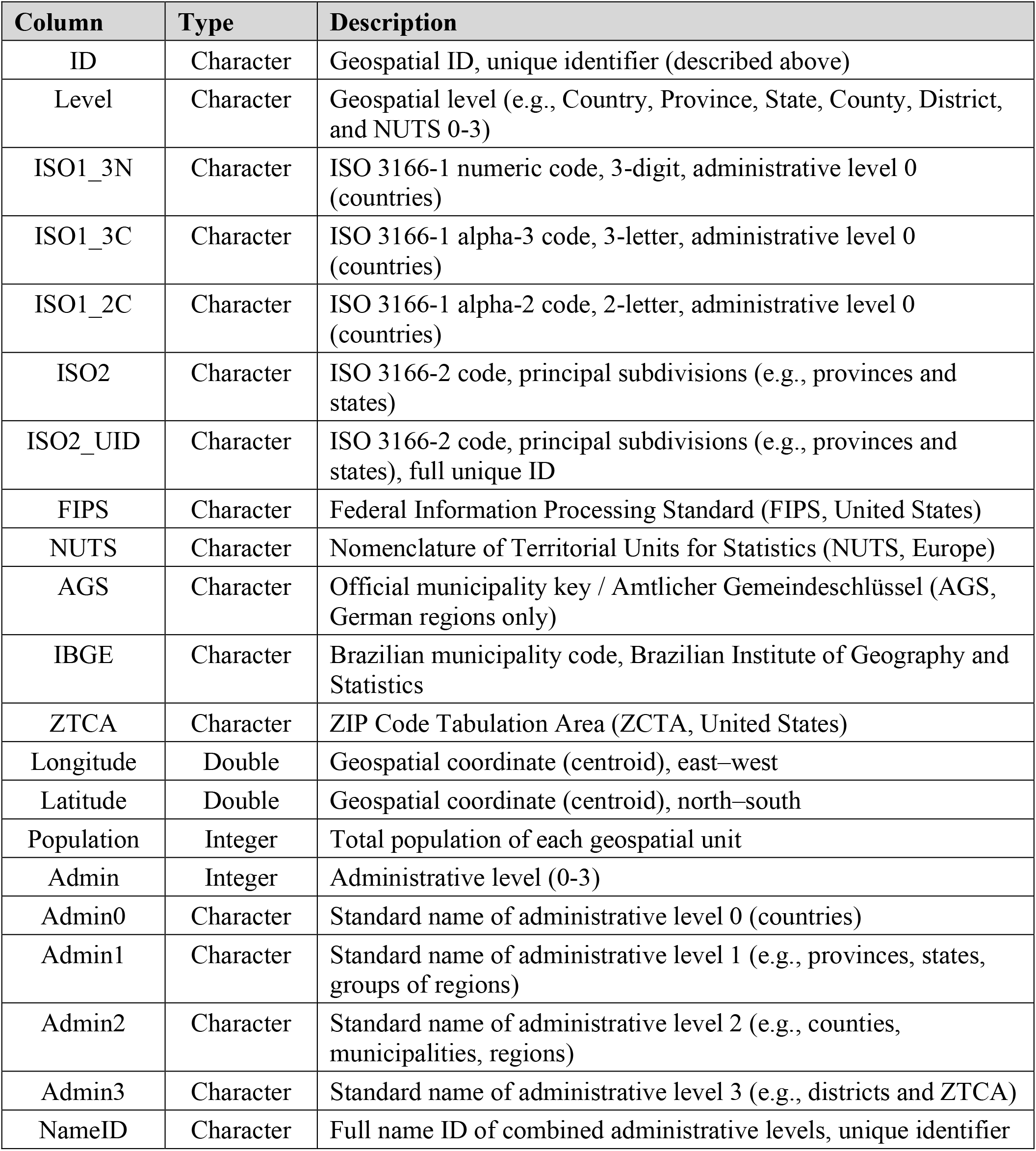
Lookup table for the unified COVID-19 dataset.

### Epidemiological Data

Daily COVID-19 case counts are taken from the different data sources, including CSSE’s JHU Coronavirus Dashboard, and georeferenced to the administrative units in which they were diagnosed.^1-12^ We merge multiple data sources with different case types. This includes translating variable names from different languages, transforming different data formats (e.g., accumulating daily counts from RKI data for Germany), and checking the aggregated counts against all data sources.

**Table 2** lists the epidemiological data structure. **Table 3** describes the different case types, including confirmed cases, deaths, hospitalizations, and testing results.

**Table 2:** COVID-19 data structure.

| Column | Type | Description |
| --- | --- | --- |
| ID | Character | Geospatial ID, unique identifier |
| Date | Date | Date of data record |
| Cases | Integer | Number of cumulative cases |
| Cases_New | Integer | Number of new daily cases |
| Type | Character | Type of the reported cases |
| Age | Character | Age group of the reported cases |
| Sex | Character | Sex/gender of the reported cases |
| Source | Character | Data source: JHU<sup>1</sup>, CTP<sup>2</sup>, NYC<sup>3</sup>, NYT<sup>4</sup>, SES<sup>5</sup>, DPC<sup>6</sup>, RKI<sup>7</sup>, JRC<sup>8</sup> |

**Table 3:** COVID-19 case types.

| Type | Description |
| --- | --- |
| Active | Active cases |
| Confirmed | Confirmed cases |
| Deaths | Deaths |
| Home_Confinement | Home confinement / isolation |
| Hospitalized | Total hospitalized cases excluding intensive care units |
| Hospitalized_Now | Currently hospitalized cases excluding intensive care units |
| Hospitalized_Sym | Symptomatic hospitalized cases excluding intensive care units |
| ICU | Total cases in intensive care units |
| ICU_Now | Currently in intensive care units |
| Negative | Negative tests |
| Pending | Pending tests |
| Positive | Positive tests, including hospitalised cases and home confinement |
| Positive_Dx | Positive cases emerged from clinical activity / diagnostics |
| Positive_Sc | Positive cases emerging from surveys and tests |
| Recovered | Recovered cases |
| Tested | Cases tested = Tests - Pending |
| Tests | Total performed tests |
| Ventilator | Total cases receiving mechanical ventilation |
| Ventilator_Now | Currently receiving mechanical ventilation |

### Hydrometeorological Data

Like many viral diseases, COVID-19 transmission is sensitive to hydrometeorological conditions, though the extent to which this impacts broad epidemiological trends has not yet been characterized. We derive meteorological variables from the second generation North American Land Data Assimilation System (NLDAS-2), using the NLDAS-2 meteorological forcings and Noah Land Surface Model simulated surface hydrological fields, and the fifth generation European Centre for Medium-Range Weather Forecasts (ECMWF) atmospheric reanalysis of the global climate (ERA5).^14,15^ Both ERA5 and NLDAS assimilate observations and model output to provide continuous maps of meteorological variables without gaps or missing values in the data, which cannot be achieved from observations alone. The fine spatial resolution of NLDAS (0.125° latitude x 0.125° longitude) and ERA5 (0.25° latitude x 0.25° longitude) represents significant improvements over earlier datasets, and both datasets have been extensively tested against observations and found to capture the observed quantities.^14,16^ ERA5 and NLDAS are available with a 4-6 day latency making these datasets particularly well-suited for forecasting COVID-19 dynamics in near real-time. NLDAS are available only for the contiguous United States, while ERA5 are available globally.

We obtain gridded hourly ERA5 and NLDAS data for January 1, 2020 onwards. Hourly data are transformed to daily mean, maximum, minimum, or total values, depending on the variable (**Table 4)**. A land-sea mask is applied to the hydrometeorological data such that any grid cells comprised of water are excluded from the analysis. Two types of average values are provided for each administrative unit: simple averages and a population-weighted averages. A small number of administrative units do not contain ERA5 or NLDAS grid cells due to their having irregular boundaries or small areal extents (e.g., ∼15% of NUTS 3 divisions). In this case, we estimate the value of meteorological values at the unit’s geographic centroid using an inverse distance weighting interpolation method and thereafter calculate the simple and population-weighted averages using these interpolated values.

**Table 4:** Hydrometeorological data structure.

| Column | Type | Unit | Description |
| --- | --- | --- | --- |
| ID | Character |  | Geospatial ID, unique identifier (described above) |
| Date | Date |  | Date of data record |
| T | Double | °C | Daily average near-surface air temperature |
| Tmax | Double | °C | Daily maximum near-surface air temperature |
| Tmin | Double | °C | Daily minimum near-surface air temperature |
| Td | Double | °C | Daily average near-surface dew point temperature |
| Tdd | Double | °C | Daily average near-surface dew point depression |
| RH | Double | % | Daily average near-surface relative humidity |
| SH | Double | kg/kg | Daily average near-surface specific humidity |
| MA | Double | % | Daily average moisture availability (NLDAS)<sup>14</sup> |
| RZSM | Double | kg/m2 | Daily average root zone soil moisture content (NLDAS)<sup>14</sup> |
| SM | Double | kg/m2 | Daily average soil moisture content (NLDAS)<sup>14</sup> |
| SM1 | Double | m3/m3 | Daily average volumetric soil water layer 1 (ERA5)<sup>15</sup> |
| SM2 | Double | m3/m3 | Daily average volumetric soil water layer 2 (ERA5)<sup>15</sup> |
| SM3 | Double | m3/m3 | Daily average volumetric soil water layer 3 (ERA5)<sup>15</sup> |
| SM4 | Double | m3/m3 | Daily average volumetric soil water layer 4 (ERA5)<sup>15</sup> |
| SP | Double | Pa | Daily average surface pressure |
| SR | Double | J/m2 | Daily average surface downward solar radiation (ERA5)<sup>15</sup> |
| SRL | Double | W/m2 | Daily average surface downward longwave radiation flux (NLDAS)<sup>14</sup> |
| SRS | Double | W/m2 | Daily average surface downward shortwave radiation flux (NLDAS)<sup>14</sup> |
| LH | Double | J/m2 | Daily average surface latent heat flux (ERA5)<sup>15</sup> |
| LHF | Double | W/m2 | Daily average surface latent heat flux (NLDAS)<sup>14</sup> |
| PE | Double | m | Daily average potential evaporation / potential latent heat flux (ERA5)<sup>15</sup> |
| PEF | Double | W/m2 | Daily average potential evaporation / potential latent heat flux (NLDAS)<sup>14</sup> |
| P | Double | mm/day | Daily total precipitation |
| U | Double | m/s | Daily average 10-m above ground Zonal wind speed |
| V | Double | m/s | Daily average 10-m above ground Meridional wind speed |
| Source | Character |  | Data source: ERA5, NLDAS ± CIESIN <sup>13-15</sup> |

**Table 4** lists the hydrometeorological variables extracted from NLDAS-2 and ERA5.

### Air Quality Data

Long-term exposure to air pollutants may increase susceptibility to severe COVID-19 outcomes.^17^ We provide long-term averages of surface-level annual average nitrogen dioxide (NO_2_) and fine particulate matter (PM_2.5_) to allow this potential impact to be incorporated into studies. We use a dataset that transforms observations of aerosol optical depth (AOD) from Earth-observing satellites to global estimates of surface-level PM_2.5_ using geophysical relationships between modeled PM_2.5_ and AOD from a chemical transport model and a Geographically Weighted Regression technique.^18^ Global NO_2_ estimates are derived by scaling the predicted concentrations from a global land use regression model with annual satellite observations of tropospheric NO_2_ columns from the Ozone Monitoring Instrument satellite.^19,20^

PM_2.5_ and NO_2_ datasets are regridded from their native resolutions (0.01° latitude x 0.01° longitude and 1 km x 1 km, respectively) to 0.05° latitude x 0.05° longitude and averaged over 2014-2018. We calculate both simple and population-weighted averages of PM_2.5_ and NO_2_ for administrative units.

### Policy Data

The time-varying policy response data described in **Table 5** are processed from the Oxford COVID-19 Government Response Tracker (OxCGRT) for the policy types listed in **Table 6**, including four categories of policies: (i) **containment and closure policies:** C1: School closing, C2: Workplace closing, C3: Cancel public events, C4: Restrictions on gatherings, C5: Close public transport, C6: Stay at home requirements, C7: Restrictions on internal movement, and C8: International travel controls, (ii) **economic policies:** E1: Income support, E2: Debt/contract relief, E3: Fiscal measures, and E4: International support, (iii) **health system policies:** H1: Public information campaigns, H2: Testing policy, H3: Contact tracing, H4: Emergency investment in healthcare, H5: Investment in vaccines, H6: Facial Coverings, H7: Vaccination Policy, and H8: Protection of elderly people, and (iv) **miscellaneous policies:** M1: Wildcard as well as policy indices for containment health, economic support, and government response. For more details, see OxCGRT’s [codebook](https://github.com/OxCGRT/covid-policy-tracker/blob/master/documentation/codebook.md), [index methodology](https://github.com/OxCGRT/covid-policy-tracker/blob/master/documentation/index_methodology.md), [interpretation guide](https://github.com/OxCGRT/covid-policy-tracker/blob/master/documentation/interpretation_guide.md), and [subnational interpretation](https://github.com/OxCGRT/covid-policy-tracker/blob/master/documentation/subnational_interpretation.md).^21^

**Table 5:** Policy data structure.

| Column | Type | Description |
| --- | --- | --- |
| ID | Character | Geospatial ID, unique identifier |
| Date | Date | Date of data record |
| PolicyType | Character | Type of the policy |
| PolicyValue | Double | Value of the policy |
| PolicyFlag | Logical | Logical flag for geographic scope |
| PolicyNotes | Character | Notes on the policy record |
| PolicySource | Character | Data source: OxCGRT<sup>21</sup> |

**Table 6:** Policy data types.

| Type | Description |
| --- | --- |
| CX | **Containment and closure policies** |
| C1 | School closing |
| C2 | Workplace closing |
| C3 | Cancel public events |
| C4 | Restrictions on gatherings |
| C5 | Close public transport |
| C6 | Stay at home requirements |
| C7 | Restrictions on internal movement |
| C8 | International travel controls |
| EX | **Economic policies** |
| E1 | Income support |
| E2 | Debt/contract relief |
| E3 | Fiscal measures |
| E4 | International support |
| HX | **Health system policies** |
| H1 | Public information campaigns |
| H2 | Testing policy |
| H3 | Contact tracing |
| H4 | Emergency investment in healthcare |
| H5 | Investment in vaccines |
| H6 | Investment in vaccines |
| H7 | Vaccination policy |
| H8 | Protection of elderly people |
| MX | **Miscellaneous policies** |
| M1 | Wildcard |
| IX | **Policy indices** |
| I1 | Containment health index |
| I2 | Economic support index |
| I3 | Government response index |
| I4 | Stringency index |
| IC | **Confirmed cases** |
| ID | **Confirmed deaths** |
| IXD | *Policy indices (Display)* |
| IXL | *Policy indices (Legacy)* |
| IXLD | *Policy indices (Legacy, Display)* |

### Other Data

#### Prevalence of Comorbid Conditions

National-level data and United States administrative level 1 data on the prevalence of underlying health conditions associated with increased risk of COVID-19 morbidity and mortality as defined by the Centers for Disease Control and Prevention (CDC) described in **Tables 7** were compiled from multiple sources. These comorbid conditions included prevalence of human immunodeficiency virus (HIV) infection, obesity, hypertension, smoking, chronic obstructive pulmonary disease (COPD), and cardiovascular disease (CVD).^22^ In addition, national-level indicators of the proportion of the population at increased risk for COVID-19 due to comorbid conditions were compiled from the estimates of Clark and colleagues and included in the unified database.^23^ Data was collected from sources online associated with reputable health organizations, health research centers, international and national organizations, research journals, and academic institutions.^23-33^ Once compiled, the final data structure was created in Microsoft Excel with all corresponding and available data.

**Table 7:** Static data structure.

| Column | Type | Unit | Description |
| --- | --- | --- | --- |
| ID | Character |  | Geospatial ID, unique identifier |
| PM2.5<sup>18</sup> | Double | µg/m<sup>3</sup> | Fine particulate matter (PM2.5) concentration (2014-2018 mean) |
| PM2.5_PopWtd<sup>18,39</sup> | Double | µg/m<sup>3</sup> | Fine particulate matter (PM2.5) concentration (2014-2018 mean, population weighted) |
| NO2<sup>19</sup> | Double | ppbv | Nitrogen dioxide (NO2) concentration (2014-2018 mean) |
| NO2_PopWtd<sup>19,39</sup> | Double | ppbv | Nitrogen dioxide (NO2) concentration (2014-2018 mean, population weighted) |
| Access_City<sup>36,37</sup> | Double | Minute | Travel time to nearest cities |
| Access_Motor<sup>38</sup> | Double | Minute | Travel time to health care facilities, with motorized transport |
| Access_Walk<sup>38</sup> | Double | Minute | Travel time to health care facilities, without motorized transport |
| Diabetes<sup>24,25</sup> | Double |  | Age-adjusted percent prevalence of adults (>=18 years old) with diagnosed diabetes |
| Obesity<sup>26-28</sup> | Double |  | Percent of obese adults (body mass index of 30+) |
| Smoking<sup>29,30</sup> | Double |  | Age-adjusted percent prevalence of adults who are current smokers |
| COPD<sup>31</sup> | Double |  | Age-standardized percent prevalence of chronic obstructive pulmonary disease by sex (Total) |
| COPD_F<sup>31</sup> | Double |  | Age-standardized percent prevalence of chronic obstructive pulmonary disease by sex (Female) |
| COPD_M<sup>31</sup> | Double |  | Age-standardized percent prevalence of chronic obstructive pulmonary disease by sex (Male) |
| CVD<sup>31</sup> | Double |  | Age-standardized percent prevalence of cardiovascular disease by sex (Total) |
| CVD_F<sup>31</sup> | Double |  | Age-standardized percent prevalence of cardiovascular disease by sex (Female) |
| CVD_M<sup>31</sup> | Double |  | Age-standardized percent prevalence of cardiovascular disease by sex (Male) |
| HIV<sup>31</sup> | Double |  | Age-standardized percent prevalence of HIV/AIDS by sex (Total) |
| HIV_F<sup>31</sup> | Double |  | Age-standardized percent prevalence of HIV/AIDS by sex (Female) |
| HIV_M<sup>31</sup> | Double |  | Age-standardized percent prevalence of HIV/AIDS by sex (Male) |
| Hypertension<sup>32,33</sup> | Double |  | Percent of adults with hypertension by sex (Total) |
| Hypertension_F<sup>33</sup> | Double |  | Percent of adults with hypertension by sex (Female) |
| Hypertension_M<sup>33</sup> | Double |  | Percent of adults with hypertension by sex (Male) |
| Risk_Tot<sup>23</sup> | Double |  | Proportion of individuals in the population that have at least 1 of the 11 identified risk conditions for COVID-19; total proportion of individuals in the population who are at any increased risk for COVID-19 |
| Risk_Age<sup>23</sup> | Double |  | Age-standardized proportion of the population that are at increased risk for COVID-19 |
| Risk_High<sup>23</sup> | Double |  | Proportion of individuals at high risk, defined as those that would require hospital admission if infected. |
| Cases_MERS<sup>34</sup> | Double |  | Total MERS cases by country (October 2012 - February 2018) |
| Cases_SARS<sup>35</sup> | Double |  | Total SARS cases by country (1 November 2002 - 7 August 2003) |
| WorldPop<sup>39</sup> | Double |  | Total population from WorldPop |
| WorldPop_Density<sup>39</sup> | Double | 1/km<sup>2</sup> | Population density from WorldPop |
| WorldPop_65<sup>39</sup> | Double |  | Population over 65 years old from WorldPop |
| WorldPop_F<sup>39</sup> | Double |  | Population by sex (Female) from WorldPop |
| WorldPop_M<sup>39</sup> | Double |  | Population by sex (Male) from WorldPop |
| Sex_Ratio<sup>39</sup> | Double |  | Sex ratio (Male / Female) from WorldPop |

#### Pandemic Preparedness

National numbers of cases from the SARS-CoV-1 and MERS outbreaks, as described in **Table 7**, were included in the unified database as proxy indicators of pandemic experience, which may be relevant for preparedness.^34,35^

#### Accessibility to cities and healthcare facilities

Population-level access to healthcare and other infrastructure may affect the trajectory of pandemics at a local scale by influencing contact rates and the introduction of new infected and susceptible individuals, as well as the speed and likelihood with which new cases are confirmed, treated, and registered in health information systems. **Table 7** lists three indicators of accessibility that are included in the unified dataset. Accessibility to nearest cities through surface transport (**Access_City**), quantified as minutes required for traveling one meter, was obtained by extracting zonal statistics from the “Accessibility to Cities 2015” raster file provided by the Malaria Atlas Project.^36^ The raster file represents the fastest traveling speed from any given point to its nearest city. It was calculated by mapping the travel time at different spatial locations and topographical conditions into grids where the fastest mode of transport took precedence.^37^ Using a similar methodology, Weiss and colleagues utilized data from OpenStreetMap, Google Maps, and academic researchers to produce maps of travel time to health care facilities with and without access to motorized transport, from which we obtained the two variables characterizing travel time (minutes) to the nearest healthcare facility by two modes of transport (**Access_Motor**: motorized transport available; **Access_Walk**: no access to motorized transport) as indicators of healthcare access.^38^

#### Population density and age structure

**Table 7** describes population density and age structure from WorldPop.^39^

Total population (**WorldPop**), population density (**WorldPop_Density**), the total population over 65 years old (**WorldPop_65**), and total population by both male (**WorldPop_M**) and female (**WorldPop_F**) were obtained by extracting zonal statistics with the 2020 unconstrained global mosaics raster files at 1km resolution from the WorldPop spatial datasets, an open access harmonized set of gridded geospatial layers with global coverage produced by drawing on census, survey, satellite and cell phone data. The ratio of male-to-female population (**Sex_Ratio**) was calculated by dividing the female population by male population.

## Data Sources

The data sources are listed in **Table 8**.

**Table 8:** Data sources of the unified COVID-19 dataset.

| Source | Description | Level |
| --- | --- | --- |
| JHU<sup>1</sup> | Johns Hopkins University Center for Systems Science and Engineering (CSSE) | Global & County/State, United States |
| CTP<sup>2</sup> | The COVID Tracking Project | State, United States |
| NYC<sup>3</sup> | New York City Department of Health and Mental Hygiene | ZCTA/Borough, New York City |
| NYT<sup>4</sup> | The New York Times | County/State, United States |
| SES<sup>5</sup> | Monitoring COVID-19 Cases and Deaths in Brazil | Municipality/State/Country , Brazil |
| DPC<sup>6</sup> | Italian Civil Protection Department | NUTS 0-3, Italy |
| RKI<sup>7</sup> | Robert Koch-Institut, Germany | NUTS 0-3, Germany |
| JRC<sup>8</sup> | Joint Research Centre | Global & NUTS 0-3, Europe |
| ERA5<sup>15</sup> | The fifth generation of ECMWF reanalysis | All levels |
| NLDAS<sup>14</sup> | North American Land Data Assimilation System | County/State, United States |
| CIESIN<sup>13</sup> | Center for International Earth Science Information Network | All levels |
| Hammer<sup>18</sup> | Global Estimates and Long-Term Trends of Fine Particulate Matter Concentrations | All levels |
| Larkin<sup>19</sup> | Global Land Use Regression Model for Nitrogen Dioxide Air Pollution; Larkin et al. (2017) | All levels |
| Anenberg<sup>20</sup> | Global surface NO2 concentrations 1990-2020 | All levels |
| OxCGRT<sup>21</sup> | Oxford COVID-19 Government Response Tracker | Global & subnational (US, UK) |
| Clark<sup>23</sup> | Lancet: Global, regional, and national estimates of the population at increased risk of severe COVID-19 due to underlying health conditions in 2020 | Global, regional, national |
| WorldPop<sup>39</sup> | WorldPop: Open Source Demographic Data and Research | All levels |
| MAP<sup>36-38</sup> | Accessibility to Cities<br>Accessibility to Healthcare | All levels |

## Technical Validation

The unified data are regularly validated before and after processing by checking and comparing all fields with the available authoritative data sources, such as the World Health Organization (WHO), the US and European Centers for Disease Control and Prevention (CDC), and between the different sources.^9-11^ Any significant discrepancy or unrealistic data (e.g., bad data fields or types, negative counts, and implausible values) are automatically detected by checking the type of the data fields (e.g. integer, double, character, or date) and rate of daily changes to investigate and correct the unified data, besides the JHU CSSE’s automatic anomaly detection system, which is designed to detect abrupt spikes or negative increases of daily cases counts. The anomaly detection and data corrections are grouped by geospatial ID, considering recent trends and total population, and data source. Moreover, the geospatial IDs are verified with the corresponding ISO codes and shapefiles for all geographic units. All components of the dataset are updated daily to sync all retrospective changes from the original sources, including any corrections or re-assignments of the case counts. The updated dataset offers more accurate and up-to-date information for researchers to model and analyze COVID-19 transmission dynamics and associations with environmental conditions.

Hydrometeorology and air quality data are all drawn from data sources that perform their own extensive evaluation routines. We did not apply additional independent evaluation of these products. Processed variables were checked for consistency with the source data to ensure that no artifacts were introduced during data transfer or spatial averaging. We perform regular checks of time-series hydrometeorological data from select administrative units in order to scan for inconsistencies or discontinuities in the ERA5 or NLDAS data records, as such errors can sometimes appear in operational Earth data products. To date we have not identified any problematic issues, but should they arise those data will be flagged as preliminary until corrected versions of the hydrometeorological data files are posted by the operational data center.

The accessibility to cities, validated by comparing it to the network distance algorithm within Google Maps, was encouraging (R^2^ = 0.66; mean absolute difference 20.7 min). The prevalence of comorbid conditions as outlined in **Tables 7** were taken from online sources directly or associated with reputable health organizations, health research centers, international and national organizations, research journals, and academic institutions. Multiple validation checks were conducted to ensure that our unified dataset matches these input sources. Pandemic preparedness data as described in **Table 7** were taken from similarly internationally-recognized research institutions and global health organizations. Multiple validation checks were conducted to ensure consistency between the unified datasets and these highly vetted data sources.

## Usage Notes

Some US counties, territories, and islands do not have standard FIPS codes or are combined from standard units such as Bristol Bay plus Lake and Peninsula Borough, Dukes and Nantucket counties, Utah jurisdictions, Federal Correctional Institution (FCI), Veterans’ Affairs, and Michigan Department of Corrections (MDOC). Those units are given a unique ID as listed in the frequently-updated lookup table on GitHub.

The Covid Tracking Project (CTP) data stopped updating on March 7, 2021, after one year of service.^2^ All other time-varying sources are currently updated/synced from the original sources on a daily basis.

The daily new cases for some units might be missing or negative when calculated from the total accumulated cases in the raw data. This can be attributed to reporting issues and reassignment of the cases. We correct and validate the data entries only when we have strong evidence to do so. Otherwise, we keep the original data exactly as obtained from the official sources. In the future, we plan to provide an augmented version of the global data at all administrative levels, derived from all data sources. Here, we maintain consistency between both the unified and raw data.

On account of the short lifetime of these pollutants and the spatial variability of their emission sources, there are sometimes substantial differences between simple and population-weighted averages, depending on the spatial distribution of the population and emission sources within administrative units.

Due to limited availability of ground monitors in some locations, the NO_2_ concentrations have greater certainty in urban areas compared with rural areas and in North America and Europe compared with other parts of the world.

The population by sex were entered as missing values for thirty-four subnational areas in Brazil since reported values were incompatible with the total population. The accessibility raster file did not cover Monaco, and the data were manually entered using values in the surrounding area. We exclude small, overseas NUTS administrative divisions (e.g., Guadeloupe, French Guiana, Réunion) from the unified dataset to decrease the computational time needed to update the dataset in near real-time. Of note, the accessibility and population data would be most relevant for analysis at subnational, rather than national level, due to the operational definition of the data.

We claim that the presentation of material therein does not imply the expression of any opinion whatsoever on the part of JHU concerning the legal status of any country, area or territory or of its authorities. The depiction and use of boundaries, geographic names and related data shown on maps and included in lists, tables, documents, and databases on this website are not warranted to be error free nor do they necessarily imply official endorsement or acceptance by JHU.

## Data Format

The data are stored in highly-compressed binary files supported by R Statistical Software and can be imported to other machine learning tools or easily converted to different formats. The R data format efficiently preserves all variable types, attributes and object classes. This offers an advantage over other common data formats, such as comma-separated values (CSV) or its compressed versions, that do not explicitly specify the variable types (e.g., integer vs double). Moreover, the produced files are much smaller in size, facilitating data access and processing.

## Data Availability

The source code used to clean, unify, aggregate, and merge the different data components from all sources will be available on GitHub.

https://github.com/CSSEGISandData/COVID-19_Unified-Dataset

## Code Availability

The source code used to clean, unify, aggregate, and merge the different data components from all sources will be available on GitHub at https://github.com/CSSEGISandData/COVID-19_Unified-Dataset.

## Acknowledgements

This work is supported by NASA Health & Air Quality project *80NSSC18K0327*, under a COVID-19 supplement, National Institute of Health (NIH) project *3U19AI135995-03S1* (“Consortium for Viral Systems Biology (CViSB)”; Collaboration with The Scripps Research Institute and UCLA), and NASA grant *80NSSC20K1122*. Johns Hopkins Applied Physics Laboratory (APL), Data Services and Esri provide professional support on designing the automatic data collection structure, and maintaining the JHU CSSE GitHub repository.

## Author Contributions

BFZ and LMG conceived and supervised the data collection and quality control. HSB created the unified dataset, standardized the administrative names and codes by geospatial ID, and harmonized the variable names and types, merged all data components, developed the main code, and is maintaining the data structure and real-time updates. BFZ and GHK processed and maintained the hydrometeorological and air quality data. All authors contributed to dataset holdings and to writing and editing the manuscript.

